# The BinaxNOW antigen detection assay on CSF samples for diagnosis of pneumococcal meningitis

**DOI:** 10.1101/2023.01.03.23284152

**Authors:** Sreeram Chandra Murthy Peela, Sujatha Sistla

## Abstract

**Introduction:** The BinaxNOW pneumococcal antigen detection assay is an immuno-chromatographic technique where the cell wall polysaccharide instead of capsular polysaccharide is the target. Although intended for use on urine samples, its role in diagnosis of pneumococcal meningitis has been reported by many others. The current study explores its accuracy when compared with real-time PCR.

**Methodology:** CSF samples from suspected meningitis cases were processed for Gram stain and culture following standard protocols. Antigen was detected by BinaxNOW antigen detection assay for *Streptococcus pneumoniae*. Previously published primers and protocols were used for real-time PCR targeting autolysin (*lyt*A) gene and was considered as the gold standard test.

**Results:** Among the 74 samples tested, pneumococcal antigen was detected in 21.6% (16/74) and qPCR in 11 (14.9%) samples. Antigen detection assay had higher sensitivity (90.9%) when compared with culture (45.5%) while the positive predictive value was the lowest (62.5% and 100% for culture). It was in substantial agreement with the gold standard test with kappa coefficient of 0.685 and an accuracy of 90.5%.

**Conclusions:** The BinaxNOW pneumococcal antigen detection test can be employed for initial screening of samples or point-of-care test owing to its substantial agreement with real-time PCR.

## Introduction

Acute pyogenic meningitis remains a major cause of morbidity and mortality in children and adults and *Streptococcus pneumoniae* is one of the three predominant etiological agents [1]. Up to 70000 deaths per year of acute bacterial meningitis were reported to be due to this organism alone in developing countries [2]. Despite its medical importance and high prevalence, diagnosis of pneumococcal meningitis may be hampered by many factors like prior antibiotic administration and time lapse before the sample is processed [3]. In developing countries like India, where over the counter availability of antibiotics is very common, it poses a challenge to the treating physician and the microbiologist to establish the etiology of acute bacterial meningitis. These difficulties were partially resolved when antigen-based assays (like latex agglutination tests or immuno-chromatographic tests) and molecular assays like PCR and real-time PCR were introduced into laboratory diagnosis. The latex agglutination tests although are rapid, they require additional equipment which may hinder their use as a point-of-care test or in resource-poor settings [3].

The BinaxNOW pneumococcal urinary antigen test is a lateral flow immuno-chromatographic test where the cell wall polysaccharide is the target instead of the capsular polysaccharide as in many latex agglutination tests. This polysaccharide is known to be similar among various pneumococcal serotypes, and hence is not influenced the serotype that may have caused the disease [4–7]. The assay requires minimal sample volume and expertise and the results can be visualised within 15 minutes. Originally it was devised to test the presence of pneumococcal antigen in urine samples of patients suspected with pneumococcal pneumonia. However, a few studies extended its use for diagnosis of pneumococcal meningitis and the test has been approved by the United States Food and Drug Administration for diagnosis of pneumococcal meningitis [3]. One advantage of this shift in testing is that the CSF samples do-not contain the antigen while a person is pneumococcal carrier (antigen test can be positive even among asymptomatic carriers). Thereby this assay can reflect the true pathological state and avoid any unintended false positives.

This prospective study was thus undertaken to assess the accuracy of the BinaxNOW antigen detection assay in comparison with real-time PCR (qPCR) for diagnosing pneumococcal meningitis. This is the second study reported from India and conducted on all age groups rather than children alone as in many other studies.

## Materials and methods

The study was conducted from June 2015 to September 2018 at a tertiary care center in southern India. The study was approved by Institute Human Ethics committee and written informed consent/assent was obtained from all study participants. CSF samples were collected from patients belonging to all age groups and clinically suspected with acute meningitis (Appendix S1). All the samples were processed for Gram stain and culture following standard procedures.

### Antigen detection

Samples were tested for pneumococcal antigen using BinaxNOW *Streptococcus pneumoniae* Antigen Card (previously Alere, Massachusetts, USA) following manufacturer instructions for performance and interpretation. A total of 35 isolates belonging to other bacterial species (*Haemophilus influenzae, Klebsiella pneumoniae, Streptococcus pyogenes, Staphylococcus aureus, Enterococcus faecalis, Enterococcus faecium* and *Escherichia coli*) and 10 pneumococcal isolates were initially tested for cross-reactivity and positivity.

### DNA extraction and qPCR

Around 180 μL of the sample was used for DNA extraction with QIAMP DNA Blood Mini kit (Qiagen, Hilden, Germany) following manufacturer protocol for Gram-positive bacteria. Real-time PCR (qPCR) targeting autolysin (*lyt*A) gene was employed for detection of pneumococci in the clinical samples using previously published primers and probe (Appendix S2) [8]. Any sample with C_T_ value ≤ 40 was considered as positive.

### Statistical analyses

The results of each test were expressed as percentages. Any sample positive by qPCR was considered as true positive. The sensitivity, specificity, positive and negative predictive values and diagnostic accuracy of culture and antigen assay in comparison with qPCR were calculated. The degree of agreement between gold standard test and other assays was estimated using Cohen’s kappa coefficient. All statistical analyses were performed in OpenEpi v3.01.

## Results

A total of 74 samples were collected during the study period among which 26 (35.1%) were females. Around 35.1% (26/74) of the subjects were children aged less than five years and 6.8% (5/74) were adults aged 60 or more years. Among the 74 subjects, qPCR was positive in 11 (14.9%) while pneumococcus was isolated from five (6.8%) patients (table 1). Four (5.4%) samples Gram stain had features suggestive of pneumococcal infection. The median C_T_ values were higher among the culture negative patients (33.13 vs. 28.76 among culture positive patients) (table 1).

**Table 1:** Results of culture and antigen tests when compared with qPCR

| Results | Culture<br>positive | Culture<br>negative | BinaxNOW<br>positive | BinaxNOW<br>negative |
| --- | --- | --- | --- | --- |
| qPCR positive | 5 (6.8 %)<sup>#</sup> | 6 (8.1%)<sup>#</sup> | 10 (13.5%)<sup>#</sup> | 1(1.4%)<sup>#</sup> |
| qPCR negative | 0 | 63 (85.1%)<sup>#</sup> | 6 (8.1%)<sup>#</sup> | 57(77%)<sup>#</sup> |
| C<sub>T</sub><sup>^</sup> | 28.8 (25,29) | 33.1 (32,34.5) | 31.2 (26.9,33.5) | 35* |
| Sensitivity<sup>@</sup> | 45.5 (21.3, 72) |  | 90.9 (62.3, 98.4) |  |
| Specificity<sup>@</sup> | 100 (94.3, 100) |  | 90.5 (80.7, 95.6) |  |
| PPV<sup>@</sup> | 100 (56.6, 100) |  | 62.5 (38.6, 81.5) |  |
| NPV<sup>@</sup> | 91.3 (82.3, 96) |  | 98.3 (90.9, 99.7) |  |
| Cohen's<br>kappa<sup>@</sup> | 0.587 (0.379 - 0.794) |  | 0.685(0.463-0.907) |  |
| Accuracy<sup>@</sup> | 91.9 (83.4, 96.2) |  | 90.5 (81.7, 95.3) |  |

\*Only one sample was antigen negative and qPCR positive; <sup>#</sup>Percentages were calculated against the total number of patients (N=74); <sup>@</sup> Estimates with 95% confidence intervals in parentheses; PPV – positive predictive value; NPV – negative predictive value; <sup>^</sup>median and interquartile ranges

The BinaxNOW test was positive when tested on all pneumococcal isolates, and negative for all other bacterial species except for one isolate of *E. faecalis*. When tested on CSF specimens, the test result was obtained in 15 minutes, and 16 (21.6%) samples were positive. Only one sample among the qPCR positive samples was negative while 6 samples negative by qPCR were positive by the antigen test. When compared with culture, the diagnostic accuracy was slightly low (91.9% for culture and 90.5% by antigen test) while sensitivity was higher (90.9% vs. 45.5% by culture) (table 1). The antigen test had substantial agreement with qPCR (Cohen’s kappa=0.685) and accuracy of 90.5% (table 1).

## Discussion

The triad of bacteria, *Streptococcus pneumoniae, Hemophilus influenzae* and *Neisseria meningitis*, are believed to be the most common pathogens causing acute pyogenic meningitis in all age groups. Etiologic diagnosis of acute pyogenic meningitis is always a challenge, as conventional microbiological techniques like Gram stain and culture are known to have poor sensitivity [9–11]. This poor sensitivity is further complicated by the antibiotic administration before sample collection. To aid in diagnosis of acute bacterial meningitis, we previously employed the latex agglutination test (Directigen™ Meningitis Combo Test Kit, Becton Dickinson, USA) on CSF samples. The assay had a sensitivity of 80% when compared with multiplex PCR where pneumolysin was the gene targeted. The assay required about 15 minutes with pre-heating steps and other equipment like centrifuge, pipettes and rotator. These requirements hindered its use as a point-of-care or a rapid test.

Antigen detection was performed using BinaxNOW antigen detection method. This is a lateral flow, immuno-chromatographic assay and detects cell wall antigen, and hence covers all the serotypes of *S. pneumoniae*. The test is rapid, simple to perform and does not require specialized equipment or training. As per the kit insert, the detection of pneumococcal antigen is maximum (>90% positive) when the number of bacterial cells is above 3 × 10^4^ cells/ml, and falls to 44% when the bacterial cell count is <1.5 × 10^4^ cells/ml. It was also mentioned that the kit had 97% specificity and 99% sensitivity. Therefore its use in the clinical diagnosis of pneumococcal meningitis has been evaluated in a few studies.

We found the test to have higher sensitivity (90.9%) but lower predictive value (62.5%) when compared with culture. In a recent multicentric study from India, 2081 CSF specimens from children suspected with bacterial meningitis were tested and highlighted the four-fold improvement in detection of pneumococcal meningitis by BinaxNOW assay (6.1% were positive by BinaxNOW test while only 1.5% by culture) [3]. When compared with culture, it had a low positive predictive value (25.6%) but had 100% sensitivity and negative predictive values. Improving on this study, we assessed the use of BinaxNOW test by comparing with qPCR, and found similar sensitivity and negative predictive values (90.9% and 98.3% respectively). Similar high rates of positivity were detected in an international study, and 19.3% were positive by culture/BinaxNOW while only 6.3% by culture/latex agglutination or 1.1% by culture alone among the Asian countries [12]. Meta-analysis by the WHO meningitis group assessed its diagnostic accuracy and found sensitivity and specificity ranging 96-100% and 95-97% respectively [3]. In a study by Picazo et al, when compared with culture or qPCR targeting autolysin gene, the sensitivity and specificity were 88% and 72.5% but both CSF and pleural fluid were tested [13].

Improving the diagnosis of pneumococcal meningitis in cases where antibiotic therapy has been initiated prior to CSF collection is one of the advantages of the BinaxNOW test. In a study by Brink et al, CSF culture was negative in all samples where antibiotic treatment preceded sample collection, while BinaxNOW assay was positive in 75% of the samples even after 10 days of therapy [14]. Similarly, in another study, the positivity by BinaxNOW test was significantly higher among patients with prior antibiotic therapy (24.2%) when compared with those without initiation of therapy (12.2%) [12]. Despite its advantages, some cross reactivity to other bacterial species has been noted. In a study on blood cultures, a weak positive reaction was identified when testing *E. faecium* and *E. faecalis* and positive results when testing *Streptococcus mitis* [15]. Similar to this, we found BinaxNOW positive for one *E. faecalis* isolate, but negative for all other pathogens tested.

The advantages of the BinaxNOW assay on CSF specimens *viz*. lack of requirement for special equipment or trained personnel, good sensitivity and specificity when compared with real-time PCR, probable indifference to prior antibiotic therapy and low turnaround time makes it a suitable method for point of care testing and especially useful in resource-limited settings. Some of the limitations of the present study are its low sample size and clinical details or histories of antibiotic therapy were not recorded.

## Conclusions

The BinaxNOW antigen detection assay can be used as point-of-care test or screening test for diagnosis of pneumococcal meningitis owing to its high accuracy, sensitivity and substantial agreement with real-time PCR.

## Data Availability

All data necessary for the manuscript has been attached in the supplementary information file. Additional data can be provided by requesting the corresponding author

## Supplementary Data

**Appendix S1: Criteria for clinical diagnosis**

**Appendix S2: Protocols**

**Table S1: Cycle threshold values**. P – Positive, N-Negative. C_T_ – cycle threshold in qPCR; Any sample with C_T_ >35 were retested (duplicates) and mean values are shown

